# Detection and Segmentation of Lesion Areas in Chest CT Scans For The Prediction of COVID-19

**DOI:** 10.1101/2020.10.23.20218461

**Authors:** Aram Ter-Sarkisov

**Affiliations:** CitAI, Artificial Intelligence Research Centre, Department of Computer Science City, University of London

## Abstract

In this paper we compare the models for the detection and segmentation of Ground Glass Opacity and Consolidation in chest CT scans. These lesion areas are often associated both with common pneumonia and COVID-19. We train a Mask R-CNN model to segment these areas with high accuracy using three approaches: merging masks for these lesions into one, deleting the mask for Consolidation, and using both masks separately. The best model achieves the mean average precision of **44**.**68**% using MS COCO criterion for instance segmentation across all accuracy thresholds. The classification model, COVID-CT-Mask-Net, which learns to predict the presence of COVID-19 vs common pneumonia vs control, achieves the **93**.**88**% COVID-19 sensitivity, **95**.**64**% overall accuracy, **95**.**06**% common pneumonia sensitivity and **96**.**91**% true negative rate on the COVIDx-CT test split (21192 CT scans) using a small fraction of the training data. We also analyze the effect of Non-Maximum Suppression of overlapping object predictions, both on the segmentation and classification accuracy. The full source code, models and pretrained weights are available on https://github.com/AlexTS1980/COVID-CT-Mask-Net.

## 1 Introduction

A number of publications showed both commonalities and differences in the manifestation of COVID-19 and common pneumonia (CP) in chest CT scans. Both conditions give rise to lesions like Ground Glass Opacity (GGO) and Consolidation (C), but they manifest in different ways. In COVID-19 patients GGO is present more often (number of lesions/scan slice), and tends to be bilateral, subsegmental C areas are also present more often compared to the patients with CP [ZYW^+^20, LFBL20]. The absolute majority of patients with COVID-19 display either GGO, or Consolidation, or a mix of both [ZZX^+^20], and GGO lesions are more diffused, larger in size, and spread over larger areas [LFBL20]. The problem with these findings is that many of them are not statistically significant, e.g. the difference in the location of lesions in [LFBL20], and sample sizes are quite small (e.g. *n* = 34 in [ZYW^+^20]). As a result, a number of machine learning methods were recently developed to assist experts in determining the diagnosis using chest CT scans.

The two-class problems (COVID-19 vs CP, COVID-19 vs Control) is inherently easier to solve due to fewer false positives than the three-class problem (COVID-19 vs CP vs Control). Some of the best solutions for the two-class problems presented in [ZZHX20, ZLS^+^20] include DenseNet169, ResNet50 and DRE-Net [SZL^+^]. Solutions for the three-class problem using chest CT scans include ResNet18 [BGCB20], ResNet50 [LQX^+^20], COVIDNet-CT [GWW20] and multiscale spatial pyramid [YWR^+^20] as feature extractors. The disadvantage of most COVID-19 detectors is either evaluating the model on a small amount of data [BGCB20, LQX^+^20], implying weak capacity for generalization, or dependence on a large dataset and data balancing tricks [GWW20, YWR^+^20] for training models.

Semantic segmentation is the prediction of object’s masks from images by predicting the class at a pixel level. Semantic segmentation models like FCN and U-Net are widely used to segment GGO, C and other lesions. These predicted masks are often used in combination with the extracted features to predict the class of the image, [ZLS^+^20, WGM^+^20], improving the final prediction over the baseline feature extractor. Models like Mask R-CNN [HGDG17] solve a the combined problem of object detection (localization) using bounding boxes and prediction of the object’s mask, known as instance segmentation. In this paper we compare three ways to predict instances of lesions’ masks. First, we use only masks for GGO areas, merging C with the background. Secondly, we merge GGO and C masks in a single ‘lesion’ mask. Finally, we keep separate masks for GGO and C instances (this approach was first presented in [TS20]). The first two are 1+1 class problem (1 object class + background, the latter is a 2+1 problem (2 object classes + background). Our choices are explained by the observations that areas with GGO have larger sizes and are observed more frequently than areas with C in COVID-19 patients, hence GGO class alone may be sufficient for COVID-19 prediction.

We show that merging GGO and C masks into one class (‘lesion’) both improves the segmentation precision and the accuracy of the classification model built on top of the segmentation model compared to using only GGO mask. We measure the model’s accuracy using MS COCO convention of Intersect over Union (IoU) thresholds [LMB^+^14]. Mask R-CNN segmentation achieves a precision of **61**.**92**%@**0**.**5** IoU, **45**.**22**%@**0**.**75** IoU and mean average precision of **44**.**68**% (across all IoU thresholds). The classifier, COVID-CT-Mask-Net [TS20] built on top of the merged masks model, achieves a COVID sensitivity of **93**.**55%** and an overall accuracy of **96**.**33%**. The classifier built on top the model with separate masks, achieves a COVID sensitivity of **93**.**88**% and an overall accuracy of **95**.**64**% on COVIDx-CT test split of the CNCB CT scans dataset. Compared to other solutions for a 3-class problem, we use only a small fraction of the dataset to get these results: 5% of the COVIDx-CT training split and 3% of the total data.

## 2 Data

The segmentation problems solved in the paper are shown in Figure 1. The 2-class problem, Figure 1b was first solved in [TS20]. We compare this problem to two 1-class problems: For the first one, Figure 1c, we only consider GGO as the positive class and train the model to detect its instances (predict the bounding box coordinates and segment the positive area within it). Consolidation (C) masks are discarded (merged with the background). For the second problem, Figure 1d, we merge the masks for GGO and C into one class (‘lesion’), thus increasing the prevalence of the positive class in the error space, compared to only GGO.

**Figure 1:**
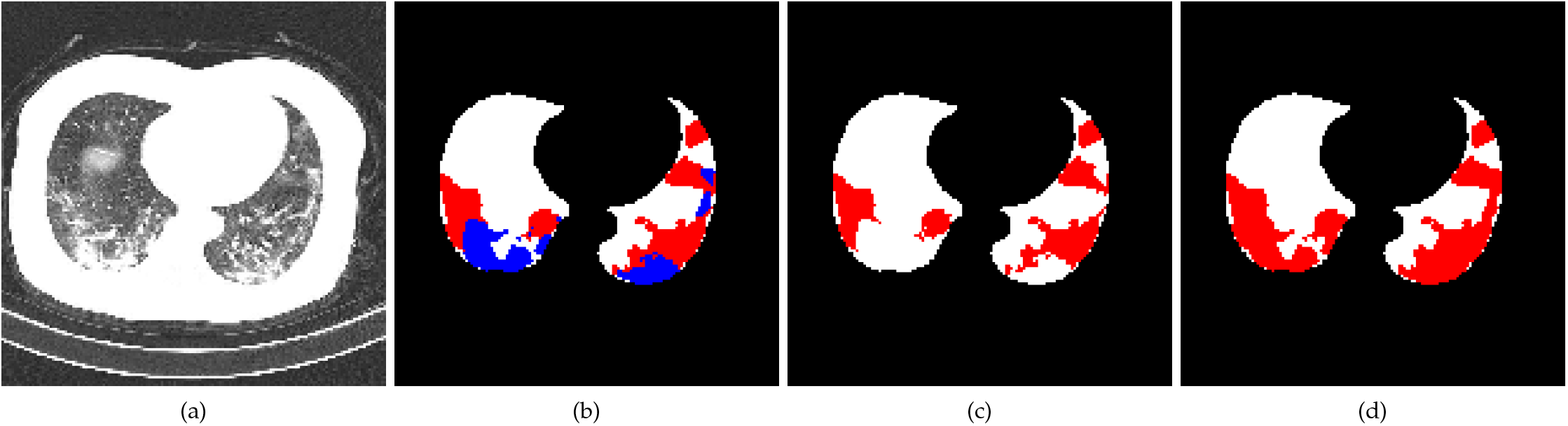
Segmentation masks for the same CT scan slice. Figure 1a: input raw image. 1b: 2-class problem, red: GGO masks, blue: C masks. figure 1c: 1-class problem (only GGO). Figure 1d: 1-class problem (merged masks for GGO and C). White masks are the lungs areas. Best viewed in color.

We use the same dataset split of 500 training + 150 validation images with varying representation of either class in each image as in [TS20]. Many images are purely negative (only background mask). To train Mask R-CNN model to solve these problems, we extract bounding box coordinates of each lesion object from the masks, and either use 3 (2 positive + 1 background label) or 2 (1 positive+1 background) labels for objects. We define each object as the area isolated from other areas of the same class either by the background or by the area of a different class. Lung mask is merged with the background for all problems. Except the usual normalization using global mean and standard deviation, no other data augmentations or balancing (resampling, class balancing, image rotations, etc) were applied to the data at any stage, unlike in many other solutions, e.g. [GWW20].

For the classification problem us re-use the train/validation/test splits in [TS20, [GWW20]. We sample 3000 images from the COVIDx-CT [GWW20] train split (1000 images/class), and use their full validation (21036 CT scans) and test (21191 CT scans) splits. As a result of our approach, we use less that 5% of the COVIDx-CT training data split, and 3% of the total CNCB CT scans data [ZLS^+^20]. Each image is the same size as in the segmentation data, 512 × 512 × 3 pixels, all alpha-channels removed. The training split used in this paper is the same as in [TS20], to have a fair comparison. As with the segmentation problem, no other data normalization tenchinques were used apart from the image global normalization.

## 3 Models and Experiments

We study in-depth the effect of non-maximum suppression (NMS) threshold, a criterion for discarding overlapping bounding box predictions in the Region Proposal Network (RPN) at train and test stages and Region of Interest (RoI) at test stage. High threshold values mean that a larger number of overlapping predictions is kept in the model. At the training stage of the segmentation model, low NMS in the RPN implies that a lower number of high-scoring predictions will be passed to RoI, and, a lower number of high-scoring predictions will be processed by RoI, both at train and test stages. This is because RoI, after passing the region of interest through the classification ‘head’ (two fully connected layers and a class+bounding box layer), can still classify this region as background, even if at the RPN stage the prediction was derived from the ‘positive’ anchor [HGDG17]. The hyperparameters of the segmentation model are set in Table 1. The model computes 4 loss functions: two by RPN (objectness and bounding box coordinates) and two by RoI (class and bounding box coordinates). For our training and evaluation we use the torchvision v0.3.0.

**Table 1:** Key hyperparameters of the segmentation models. RPN output is the number of predictions after the NMS step, RoI output is the maximum number of predictions at test stage after the NMS stage, RPN score_*θ*_ is the threshold for positive predictions at train time, RoI score_*θ*_ is the threshold for object confidence at test time. In RoI, NMS threshold is used only in testing.

| Backbone | Anchor sizes | Anchor scales | RPN NMS$_{\theta}$ | RoI NMS$_{\theta}$ | RPN sample | RoI sample | RPN Output | RoI Output | RPN score$_{\theta}$ | RoI score$_{\theta}$ |
| --- | --- | --- | --- | --- | --- | --- | --- | --- | --- | --- |
| ResNet50 +FPN | 2<sup>2:5</sup> | 0.1, 0.25, 0.5, 1, 1.5, 2 | 0.25/0.75 | -<br>0.25/0.75 | 256<br>- | 256<br>- | 1000 | 128 | 0.75<br>- | -<br>0.75 |

In COVID-CT-Mask-Net, see Figures 2 and 3, Mask R-CNN layers, including RPN and RoI are set to test mode: they don’t compute any losses. Therefore, RoI uses NMS threshold to filter predictions. A larger number of overlapping positive prediction can prompt the model to learn to associate them with a particular class, e.g. they are more prevalent in COVID-19 rather than common pneumonia. If the NMS threshold is low, the model will have to learn to associate a small number of distant predictions with the particular condition, which is likely to be a harder problem, because of the similarities between COVID-19 and common pneumonia. RoI score_*θ*_ is set to − 0.01 to accept all predictions regardless of confidence score, to keep the input size in the classification module **S** of fixed size. The details of the architecture of the classification model (including the batch to feature conversion) are presented in Figures 2 and 3 and [TS20], and its hyperparameters in Table 2.

**Table 2:** Key hyperparameters of COVID-CT-Mask-Net. Backbone network, anchor scales and sizes are the same as in Table 1. Both RPN and RoI modules are set to the test mode. RoI score_*θ*_ is set to − 0.01 to accept all predictions, even with low scores, to maintain the fixed batch size that is passed to the classification module **S**. The value of **S** is the total number of trainable parameters in it.

| Backbone | Anchor sizes | Anchor scales | RPN $NMS_{\theta}$ | RoI $NMS_{\theta}$ | RPN Output | RoI output/ Batch size | RPN $score_{\theta}$ | RoI $score_{\theta}$ | Classifier Module **S** |
| --- | --- | --- | --- | --- | --- | --- | --- | --- | --- |
| ResNet50 +FPN | $2^{2:5}$ | 0.1, 0.25, 0.5, 1, 1.5, 2 | 0.25/0.75 | 0.25/0.75 | 1000 | 256 | - | - 0.01 | 2.26M |

**Figure 2:**
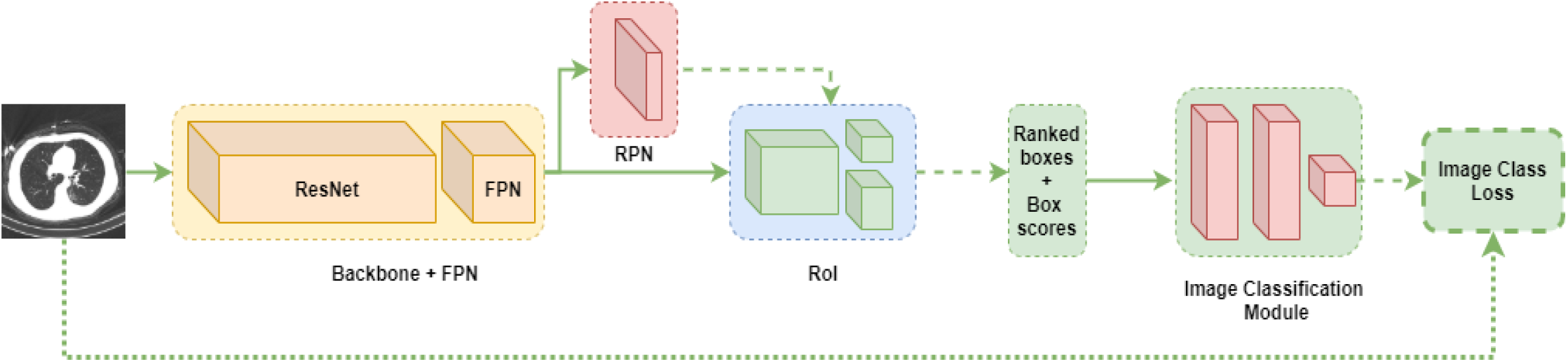
Architecture of the COVID-CT-Mask-Net classification model.

**Figure 3:**
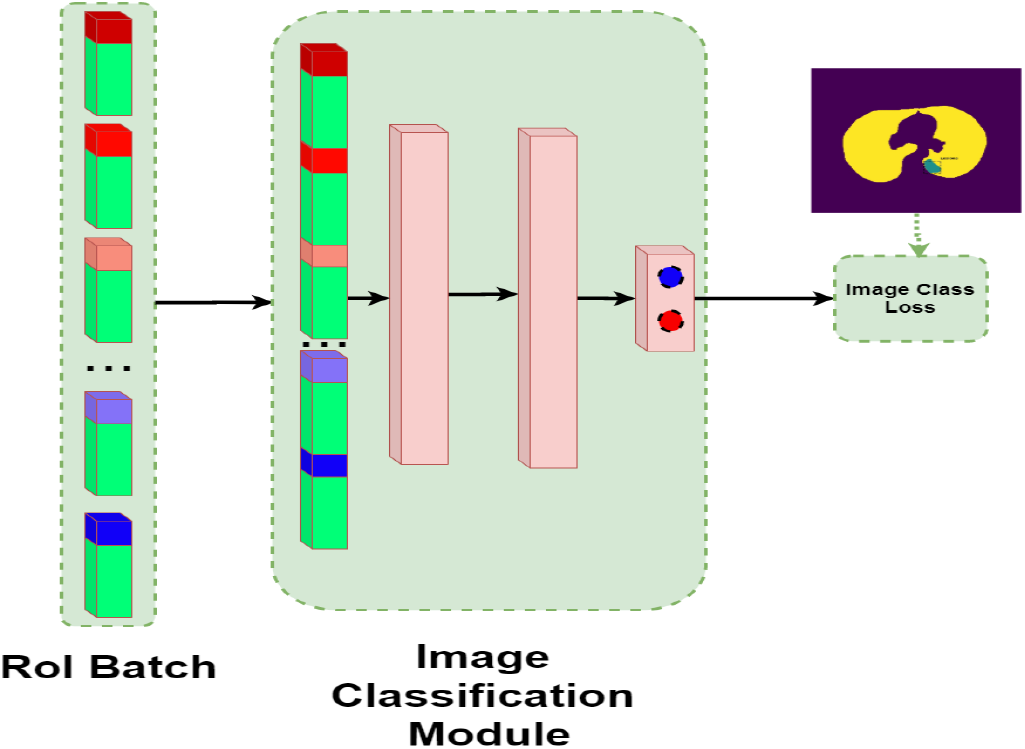
Architecture of the Image Classification Module **S**.

### 3.1 Experimental results

Each segmentation model was trained using Adam optimzier with the same learning rate of 1*e* − 5 and weight regularization coefficient 1*e* − 3 for 100 epochs, the best models for each configuration are reported in Table 4. Training of each model took about 3h on a GPU with 8Gb VRAM. All classifiers were trained with the same configuration for 50 epochs, which took about 8 hours on the same GPU. The sizes of the models are presented in Table 3, the difference in size between all segmentation models presented here is minuscule (< 1000 parameters). The architecture and the training of models with separate masks is exactly the same as in [TS20], the only difference, that explains better results in Tables 4-6 is due to the removal of very small objects (less than 10 × 10 pixels) and reduction of unnecessary large sample sizes during the training of RPN and RoI, from 1024/1024 in [TS20] to 256/256 in this paper.

**Table 3:**
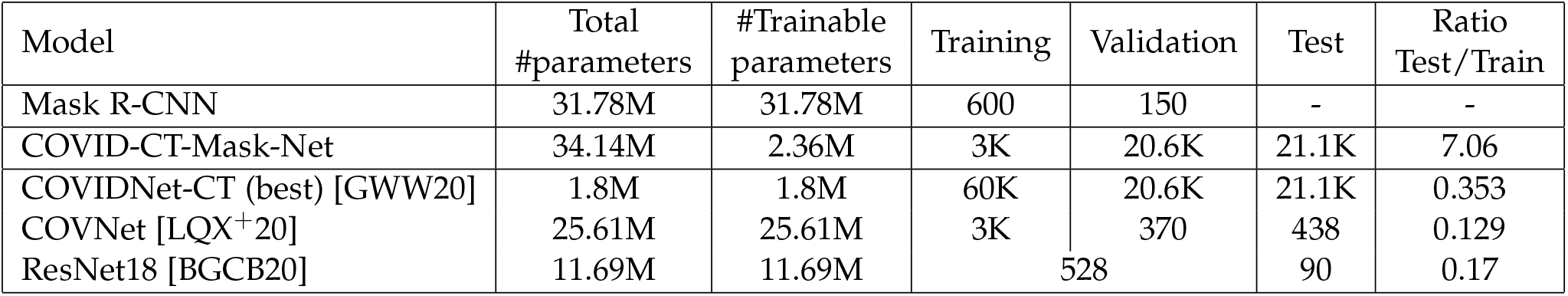
Comparison of the models’ sizes and data splits used to training, validation and testing. The number of the trainable parameters in COVID-CT-Mask-Net is due to the fact that we only train the module **S** and batch normalization layers in the backbone.

| Model | Total #parameters | #Trainable parameters | Training | Validation | Test | Ratio Test/Train |
| --- | --- | --- | --- | --- | --- | --- |
| Mask R-CNN | 31.78M | 31.78M | 600 | 150 | - | - |
| COVID-CT-Mask-Net | 34.14M | 2.36M | 3K | 20.6K | 21.1K | 7.06 |
| COVIDNet-CT (best) [GWW20] | 1.8M | 1.8M | 60K | 20.6K | 21.1K | 0.353 |
| COVNet [LQX<sup>+</sup>20] | 25.61M | 25.61M | 3K | 370 | 438 | 0.129 |
| ResNet18 [BGCB20] | 11.69M | 11.69M | 528 |  | 90 | 0.17 |

**Table 4:** Average precision of segmentation models.

| Model | AP@0.5 | AP@0.75 | AP@[0.5 : 0.95] |
| --- | --- | --- | --- |
| Only GGO mask + NMS@0.25 | 0.4575 | 0.3777 | 0.3542 |
| Only GGO mask + NMS@0.75 | 0.4588 | 0.3982 | 0.3610 |
| Merged masks + NMS@0.25 | 0.5682 | 0.4134 | 0.4310 |
| Merged masks + NMS@0.75 | **0.6192** | **0.4522** | **0.4468** |
| Separate masks + NMS@0.25[TS20] | 0.4741 | 0.3895 | 0.3641 |
| Separate masks + NMS@0.75[TS20] | 0.5020 | 0.4198 | 0.3871 |

To measure the accuracy of the segmentation models, we use the average precision (AP), a benchmark tool for datasets labelled at an instance level like MS COCO[LMB^+^14] and Pascal VOC [EVGW^+^10]. We adapt the MS COCO convention and report values for three thresholds: AP@0.5, AP@0.75 and AP (primary challenge metric). The first two use Intersect over Union (IoU) between predicted and ground truth bounding boxes with thresholds equal to 0.5 and 0.75. The latter averages over thresholds between 0.5 and 0.95 with a 0.05 step (a total of 10 thresholds). For details see [LMB^+^14]. We adapt the implementation of average precision computation from https://github.com/matterport/Mask_RCNN. Confidence threshold for considering the object (RoI_*θ*_ hyperparameter) is 0.75 across all models. Only predictions with confidence scores >RoI_*θ*_ are considered for computing (m)AP, the rest are discarded. RoI NMS_*θ*_ is always the same as RPN.

Images in Figure 4 illustrate the difference between the two NMS thresholds across each all mask types. Each column corresponds to a particular CT scan slice. The bottom row is the ground truth masks with both segmented lesion regions. Rows 1,3,5 are models that use NMS threshold of 0.25, rows 2,4,6 use NMS threshold of 0.75. Rows 1,2 are models that were trained only with the GGO mask. Models in rows 3,4 were trained with merged masks. Models in rows 5,6 were trained using both masks. Models with a higher NMS threshold output a larger number of predictions overall (except, for example, in Figure 4e, the models with the merged GGO and C masks, row 3 with low NMS and row 4 with high NMS), many of them overlapping. This is a consequence of the fact that a particular predicted region can have a high enough confidence score in RPN to be passed on to RoI, but then RoI classification ‘head’ outputs a confidence score lower than RoI score_*θ*_, hence that region will be classified as background. In case of a low NMS, an overlapping prediction with a slightly lower score would be discarded at RPN stage. In case of the high NMS, it would be added to the pool of predictions, and RoI could extract a confidence score exceeding RoI score_*θ*_ from this second prediction, therefore, models with high NMS produce more predictions overall, both true and false positives.

**Figure 4:**
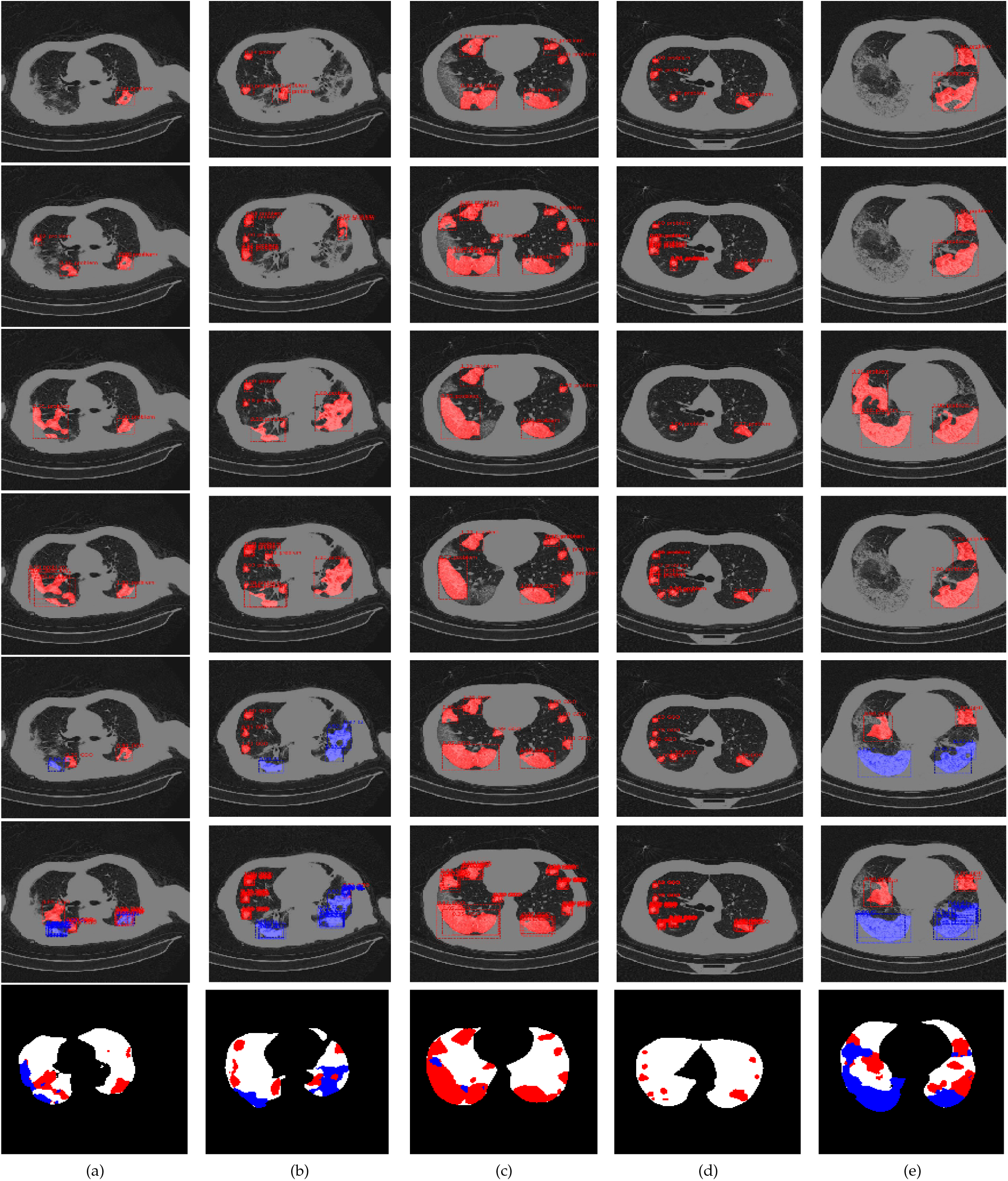
Predicted masks for a number of CT scans. Row 7: ground truth masks, red: GGO, blue: C. Rows 1,3,5: models with NMS=0.25. Rows 2,4,6: models with NMS=0.75. Rows 1,2: models trained only with the GGO mask, Rows 3,4: models trained with the merged GGO and C masks. Rows 5,6: models trained with separate masks for both classes. All mask predictions are overlaid with bounding boxes and RoI confidence scores. Best viewed in color.

Evaluation results of the segmentation model are summarized in Table 4. Models using high NMS threshold of 0.75 outperform the ones with low NMS threshold of 0.25 across all mask types. The model that learns from merged GGO and C masks with high NMS confidently outperforms GGO-only at every level of the IoU threshold. Apart from the NMS effect described above, GGO and C areas in CT scans have many commonalities, so if the model learns to segment GGO only, then Consolidation and background have the same label. As a result, the model associates some important patterns with the background rather than the object class. Results for separate GGO and C masks are mostly better than for only GGO, but worse than for the merged masks. We explain this by the fact that overall C is not very well represented in the dataset (see [TS20] for details of the data analysis), and therefore the model often confuses it with GGO features, or fails to learn certain important features because of their under-representation in the data.

Results of the COVID-CT-Mask-Net evaluation are presented in Table 5, and the comparison of the best models we trained (highest COVID sensitivity and highest overall accuracy) in Table 6. All results are a significant improvement over the baseline COVID-CT-Mask-Net model in [TS20], which we beat by 3.08% (COVID sensitivity) and 5.10% (overall accuracy). Comparing the segmentation and classification results though, the advantage of the segmentation models learning from merged masks doesn’t immediately translate into the advantage for solving the classification problem. Overall, the classifiers derived from these models are slightly better than the classifiers derived from the segmentation models for two classes, and noticeably better than GGO-only models. This advantage, though is much smaller than than the gap in the AP and mAP metrics for the corresponding segmentation problems.

**Table 5:** Sensitivity (precision) and overall accuracy results on COVIDx-CT test data (21192 images). Best results in bold.

| Model | COVID-19 | Pneumonia | Normal | Overall |
| --- | --- | --- | --- | --- |
| Only GGO mask + NMS@0.25 | 93.39% (95.73%) | 95.27% (93.08%) | 97.30% (97.95%) | 95.77% |
| Only GGO mask + NMS@0.75 | 86.98% (92.26%) | 94.27% (69.70%) | 71.12% (94.75%) | 82.45% |
| Merged masks + NMS@0.25 | 93.56% (97.92%) | **97.20%** (90.99%) | 95.12% (98.34%) | 95.52% |
| Merged masks + NMS@0.75 | 92.68% (96.29%) | 96.69% (93.63%) | **97.74%** (98.54%) | **96.33%** |
| Separate masks + NMS@0.25 | 92.22% (95.51%) | 93.06% (90.11%) | 95.15% (96.08%) | 93.82% |
| Separate masks + NMS@0.75 | **93.88%** (95.88%) | 95.06% (93.00%) | 96.91% (97.66%) | 95.64% |
| COVID-CT-Mask-Net (best) [TS20] | 90.80% (94.75%) | 91.62% (87.08%) | 91.10% (94.33%) | 91.66% |

**Table 6:**
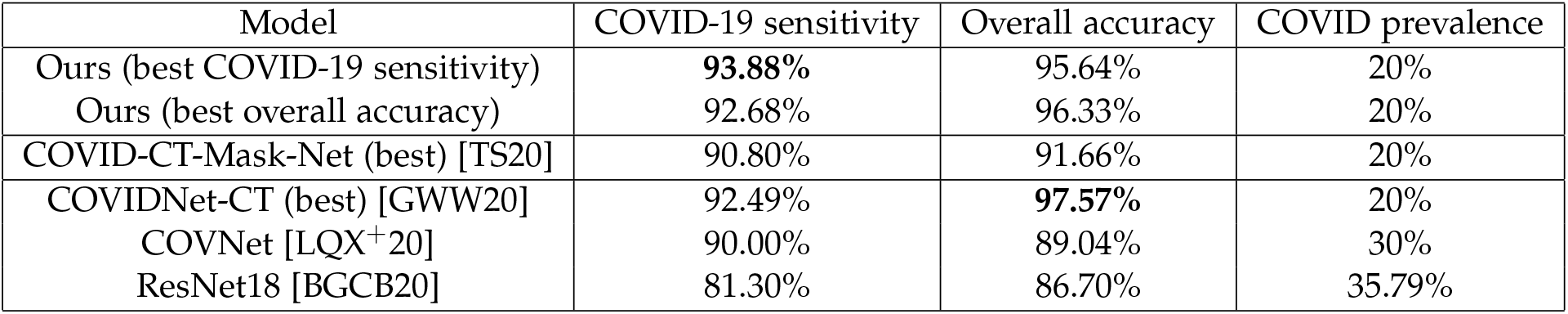
Comparison to other models. The results for COVIDNet-CT were obtained by running the publicly available model (https://github.com/haydengunraj/COVIDNet-CT) on the same test split using inference method, results for the other two models are taken from the publication. Last column is the share of COVID observations in the test split. Test split for COVNet has 438 images, ResNet18 90 images.

Compared to benchmark models, we beat COVIDNet-CT [GWW20] by 1.07% in COVID-19 sensitivity.

## 4 Conclusions

In this paper we compared a number of Mask R-CNN models that detect and segment instances of two types of lesions in chest CT scans. We established that merging lesion masks for Ground Glass Opacity and Consolidation into a single lesion mask greatly improves the predictive power and the precision of the instance segmentation model compared to other approaches. We extended these model to predict COVID-19, common pneumonia and control classes using COVID-CT-Mask-Net architecture. On a large COVIDx-CT dataset (21192 chest CT scan slices), the classification model derived from the best segmentation model achieved the COVID-19 sensitivity of 92.68% and an overall accuracy of 96.33%, and the model derived from the segmentation model using both masks achieved a COVID-19 sensitivity of 93.88% and an overall accuracy of 95.64%. The source code and the pretrained models are available on https://github.com/AlexTS1980/COVID-CT-Mask-Net.

## Data Availability

All data, algorithms, code, pretrained weights, etc are publicly available.

https://github.com/AlexTS1980/COVID-CT-Mask-Net

